# Total Beating-Heart Aortic Arch Repair Without Cardiac Arrest: A Proof-of-Concept Study

**DOI:** 10.64898/2026.05.28.26354390

**Authors:** K. Wisniewski, AM. Dell’ Aquila, V. Carranza Porras, F. Dinkel, S. Martens, A. Rukosujew

**Affiliations:** Department of Cardiothoracic Surgery, University Hospital Münster, Münster, Germany; Department of Cardiac Surgery, University Hospital Halle (Saale), Halle (Saale), Germany

**Author notes:** **Corresponding Author:** Konrad Wisniewski, MD, Department of Cardiothoracic Surgery, University Hospital Münster, Albert-Schweitzer-Campus 1, 48149 Münster.

**Keywords:** aortic arch surgery, beating heart, myocardial protection, continuous myocardial perfusion, frozen elephant trunk

## Abstract

**Background:** Cardioplegic arrest during complex aortic arch repair imposes prolonged global myocardial ischaemia, which may contribute to postoperative low cardiac output syndrome (LCOS) and mortality. Whether cardioplegic arrest can be entirely avoided — performing the complete procedure on a continuously perfused, beating heart — has not previously been evaluated in a clinical series.

**Methods and Results:** Between November 2017 and January 2026, 29 consecutive patients underwent total beating-heart aortic arch repair without any cardioplegic arrest at a single centre. Continuous antegrade myocardial perfusion (warm blood, 34°C, 300–400 mL/min, perfusion pressure 60–80 mmHg) was delivered via an aortic root needle vent throughout each procedure. Two variants were employed: axillary cannulation with selective antegrade cerebral perfusion (n = 24, 82.8%), and direct aortic cannulation with extra-anatomical left carotid bypass for distal Zone 2 pathology (n = 5, 17.2%). Mean age was 55.4 ± 13.6 years; 41.4% presented with aortic dissection (B/non-A-non-B). No patient required conversion to cardioplegic arrest. Perioperative myocardial infarction and LCOS occurred in none of the patients. Median peak CK-MB was 44.0 U/L. Thirty-day mortality was 10.3% (n = 3); all deaths were due to respiratory failure or visceral ischaemia complicating acute type B dissection.

**Conclusions:** Total beating-heart aortic arch repair without cardioplegic arrest is technically feasible and clinically safe in appropriately selected patients and is associated with the complete absence of perioperative myocardial infarction and LCOS across a heterogeneous, high-risk cohort. These findings support prospective, multicentre evaluation of no-arrest myocardial protection as a strategy to reduce the cardiac morbidity of complex arch surgery.

## Introduction

Aortic arch surgery has undergone a fundamental transformation over the past three decades. The introduction of branched prostheses, the frozen elephant trunk (FET) technique, and refined cerebral perfusion strategies have collectively expanded the boundaries of resectability in arch reconstruction to levels unimaginable in the era of deep hypothermic circulatory arrest.(1,2). Organ protection in aortic arch surgery has markedly improved over the past decades. The challenge of cerebral protection, which dominated the technical discourse of aortic arch surgery for decades, has been substantially addressed(3,4).

What has received comparatively little attention is the myocardium. In most contemporary arch operations, the heart is arrested by cardioplegic infusion and maintained in global ischaemia for the entire duration of the procedure. In complex cases involving concomitant aortic root replacement, valve surgery, or coronary revascularisation, this period of global myocardial ischaemia frequently exceeds 90 to 120 minutes, and in some cases 150 minutes or more(5,6). Even with intermittent cold blood cardioplegia administered at regular intervals, such arrest durations impose a cumulative ischaemia-reperfusion burden on the myocardium that is without parallel in any other domain of cardiac surgery.

The clinical consequence is well known: a substantial proportion of patients undergoing complex arch repair may develop postoperative low cardiac output syndrome (LCOS), requiring prolonged inotropic support and representing the initiating event in a cascade that culminates, in its most severe form, in multiorgan dysfunction and death.

The concept of continuous non-cardioplegic myocardial perfusion during arch repair — maintaining coronary blood flow throughout the procedure rather than interrupting it — was first described before and was developed into a systematic clinical strategy by Martens and colleagues, who demonstrated that transitioning from cardioplegic arrest to continuous myocardial perfusion at the completion of cardiac procedures significantly reduced cardiac ischaemia time, LCOS, and operative mortality(7,8). Their work established the feasibility of operating on the aortic arch with a perfused, beating heart. This technique has been adopted in multiple centres — including ours.

However, in every previously reported series employing continuous myocardial perfusion, the technique was initiated after an initial period of cardioplegic arrest. The question of whether global myocardial ischaemia can be eliminated entirely from arch surgery has not previously been answered in a clinical series.

The present study reports the first institutional experience with a strategy of total beating-heart aortic arch repair without any cardioplegic arrest, in which continuous antegrade myocardial perfusion is established at the beginning of the procedure and maintained uninterrupted throughout arch reconstruction. We describe the technical evolution of the approach across two operative variants, report outcomes in 29 consecutive patients and assess the feasibility and safety of this strategy as a proof of concept for a fundamentally different paradigm of myocardial protection in complex arch surgery.

## Methods

### Study Design and Patient Selection

This was a single-centre, retrospective, proof-of-concept study conducted at the Department of Cardiothoracic Surgery, University Hospital Münster, Germany. All patients who underwent complex aortic arch repair using a total beating-heart technique — defined as continuous antegrade myocardial perfusion throughout the entire procedure without any period of cardioplegic arrest — between November 2017 and January 2026 were included. The study was approved by the institutional ethics committee, and the requirement for individual informed consent was waived given the retrospective nature of the analysis.

The beating-heart approach was applied in patients fulfilling the following anatomical prerequisites: a macroscopically normal aortic root and ascending aorta and absence of significant coronary artery disease, no pre-existing severe left ventricular dysfunction. In the early phase of the programme, the technique was also applied in selected cases of acute aortic dissection (Type B, non-A-non-B); however, as institutional experience accumulated, number of patients with acute dissection was progressively reduced owing to the fragility of the dissected aortic wall. Additionally, two patients in the early series underwent concomitant right coronary artery bypass grafting after unexpected right ventricular dysfunction was identified upon separation from cardiopulmonary bypass; both had pre-existing in-stent lesions of the right coronary artery that had been deemed haemodynamically insignificant on preoperative evaluation. Following these cases, the presence of any coronary artery disease was adopted as an exclusion criterion for this technique. All procedures were performed by a single surgeon (A.R.).

### Surgical Technique

All operations were performed through a median sternotomy. Two technical variants were employed, selected according to the anatomical extent of the aortic pathology and the planned distal reconstruction zone.

### Variant 1: Axillary Cannulation with Needle-Vent Myocardial Perfusion (Zones 0–1)

In 24 patients (82.8%) with pathology involving Zones 0–1 of the aortic arch, cardiopulmonary bypass (CPB) was established via cannulation of the right axillary artery for arterial inflow and the right atrium for venous drainage (Figure 1). After initiation of CPB, systemic cooling to a target nadir oesophageal temperature of 28–30°C was commenced.

**Figure 1.**
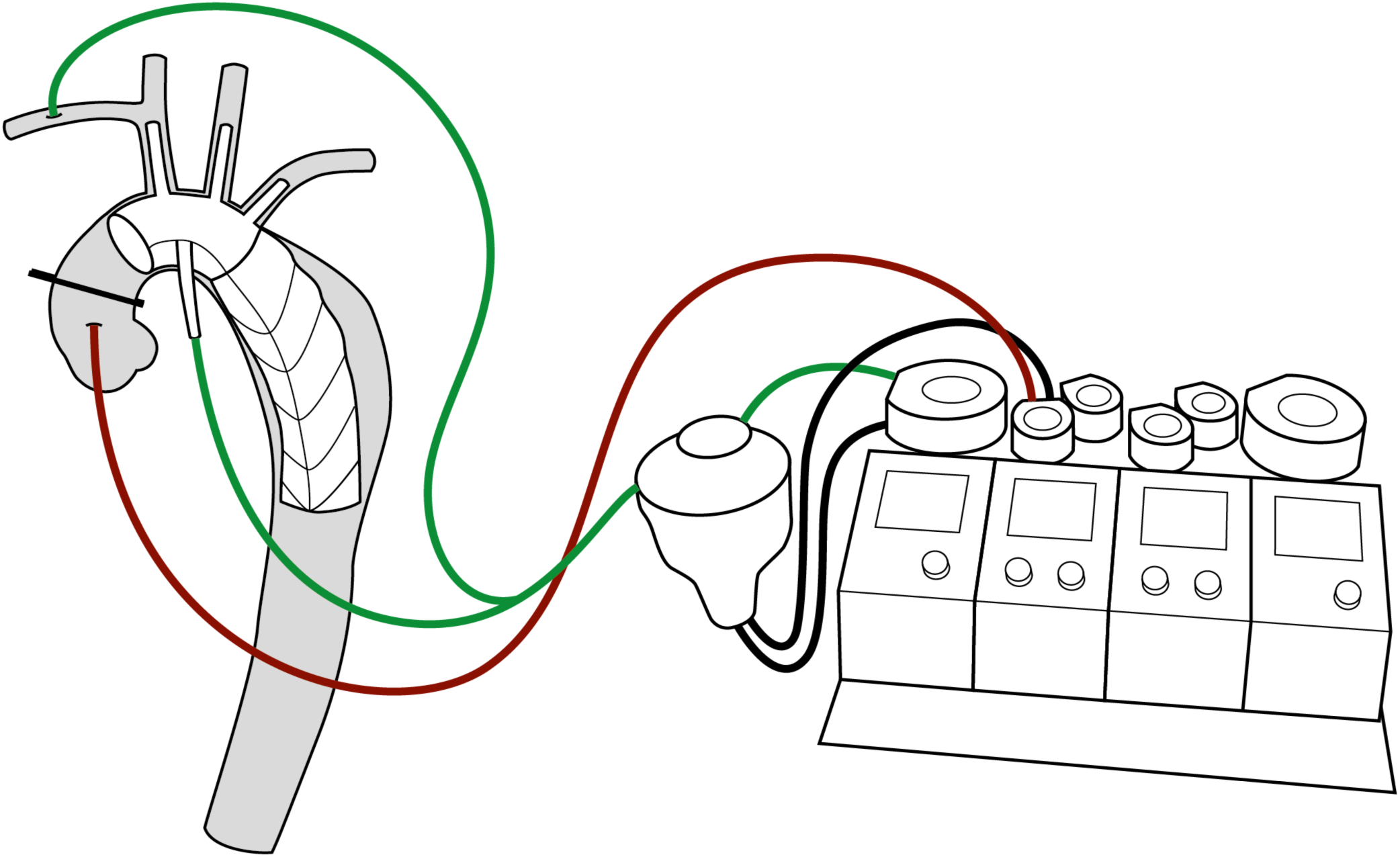
Perfusion circuit schematic for Variant 1 (axillary cannulation with needle-vent myocardial perfusion). Systemic and cerebral perfusion is delivered via the right axillary artery (green line), providing the route for selective antegrade cerebral perfusion (SACP) during circulatory arrest. A dedicated side-branch from the cardiopulmonary bypass circuit supplies continuous antegrade myocardial perfusion via a needle vent inserted into the aortic root (red line), delivering warm oxygenated blood (34°C) at a flow of 300–400 mL/min and a target perfusion pressure of 60–80 mmHg. (SACP = selective antegrade cerebral perfusion)

A 12-gauge needle vent (normally used for administration of cardioplegia) was then inserted directly into the intact aortic root and connected to the separate pump of the CPB circuit via a dedicated side-branch line. The ascending aorta was cross-clamped at the sinotubular junction (proximal clamp), isolating the aortic root for myocardial perfusion. This established continuous antegrade myocardial perfusion with warm oxygenated blood (34°C) at a flow of 300–400 mL/min. Myocardial perfusion pressure was monitored continuously via the needle-vent line and maintained at a target of 60–80 mmHg through adjustment of separate pump flow. Left ventricular decompression was maintained throughout via a vent placed in the right superior pulmonary vein. No cardioplegic solution was administered at any point during the procedure (Figure 2).

**Figure 2.**
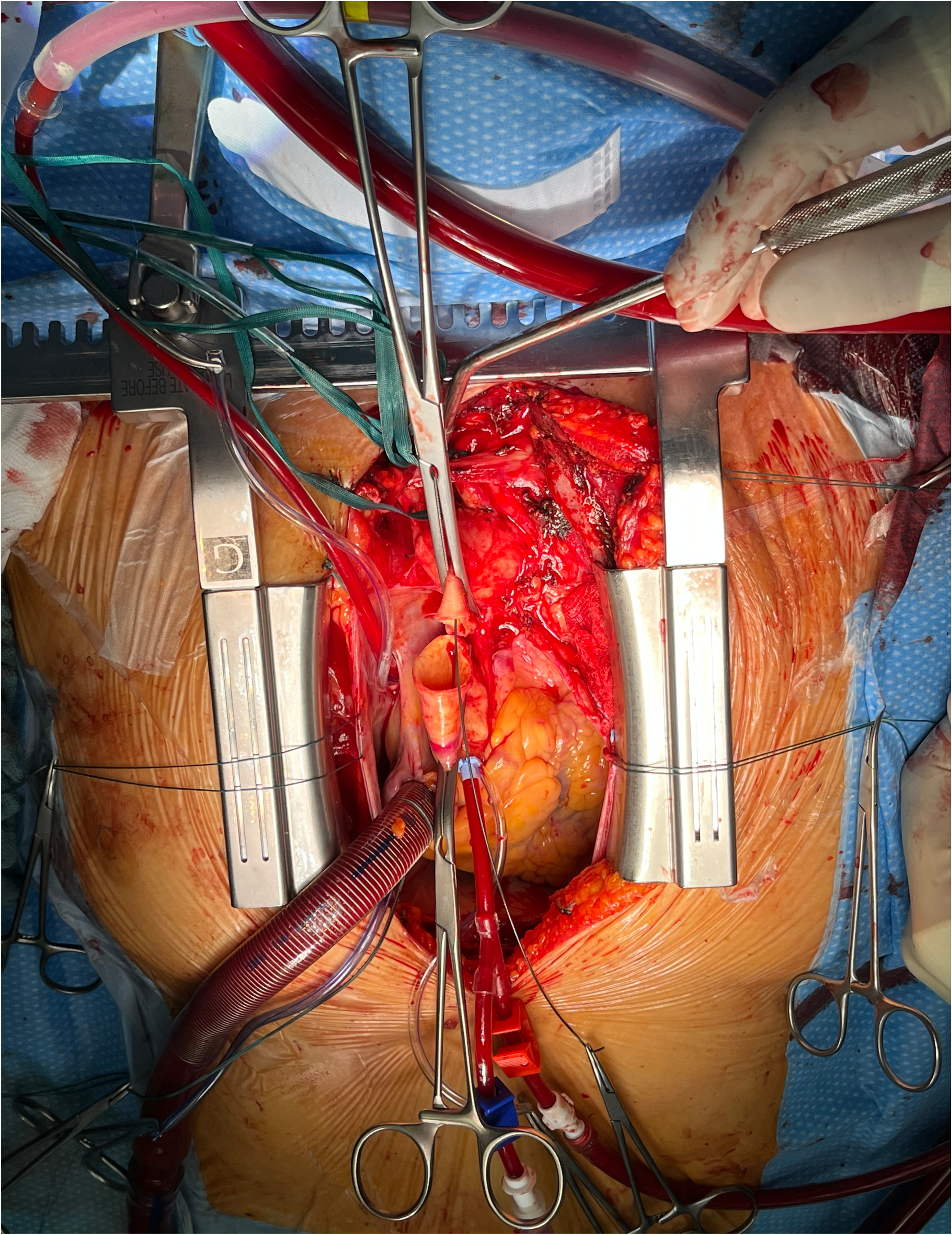
Intraoperative photograph illustrating Variant 1: needle-vent continuous myocardial perfusion. The ascending aorta is cross-clamped at two levels: immediately proximal to the innominate artery (distal clamp) and at the sinotubular junction (proximal clamp), isolating the aortic root for myocardial perfusion. A 12-gauge needle vent is inserted directly into the aortic root and connected to the dedicated myocardial perfusion line (red tubing), through which continuous antegrade warm-blood perfusion is delivered at controlled pressure. The perfusion line is visible in the operative field. The heart continues to beat throughout, with no cardioplegic arrest applied at any stage of the procedure.

Once the target core temperature was reached, lower-body flow was reduced, the ascending aorta was cross-clamped immediately proximal to the innominate artery (distal clamp) and the aortic arch was opened. Selective antegrade cerebral perfusion (SACP) was delivered via the right axillary artery with a perfusion target of 80 - 90 mmHg. Bilateral cerebral oximetry was monitored continuously using near-infrared spectroscopy (NIRS) in all patients throughout the procedure. Perfusion catheter was selectively inserted into the left common carotid artery only if a clinically significant reduction in NIRS signal was observed; prophylactic bilateral cannulation was not performed as a routine. Arch reconstruction — total/subtotal replacement, with or without frozen elephant trunk (FET) implantation — was then carried out during short circulatory arrest, while continuous antegrade myocardial perfusion was maintained.

The heart remained beating throughout the entire procedure. No attempt was made to induce asystole. As a possible bailout strategy in case of ST-segment elevation or other signs of myocardial ischemia, cardioplegia would be administered. In case of ventricular fibrillation, defibrillation would be performed. In our series of cases, none were needed.

On completion of the distal anastomosis, and lower-body perfusion was re-established via the sidearm of the prosthesis. Rewarming proceeded in standard fashion. Anastomosis of the aortic arch prosthesis and aortic root was performed. The myocardial perfusion needle was withdrawn, and the proximal aortic clamp was released. Then the reconstruction of the supra-aortic vessels was then completed, followed by weaning from CPB (Video 1 and 2).

### Variant 2: Direct Aortic Cannulation with Extra-Anatomical Left Carotid Bypass (Zone 2)

In five patients (17.2%) with pathology extending to distal Zone 2 of the aortic arch in whom the proximal ascending aorta remained anatomically suitable for direct cannulation, a modified perfusion strategy was employed to provide simultaneous myocardial and bilateral cerebral perfusion from a single arterial access site. CPB was established via direct cannulation of the proximal ascending aorta (Figure 3).

**Figure 3.**
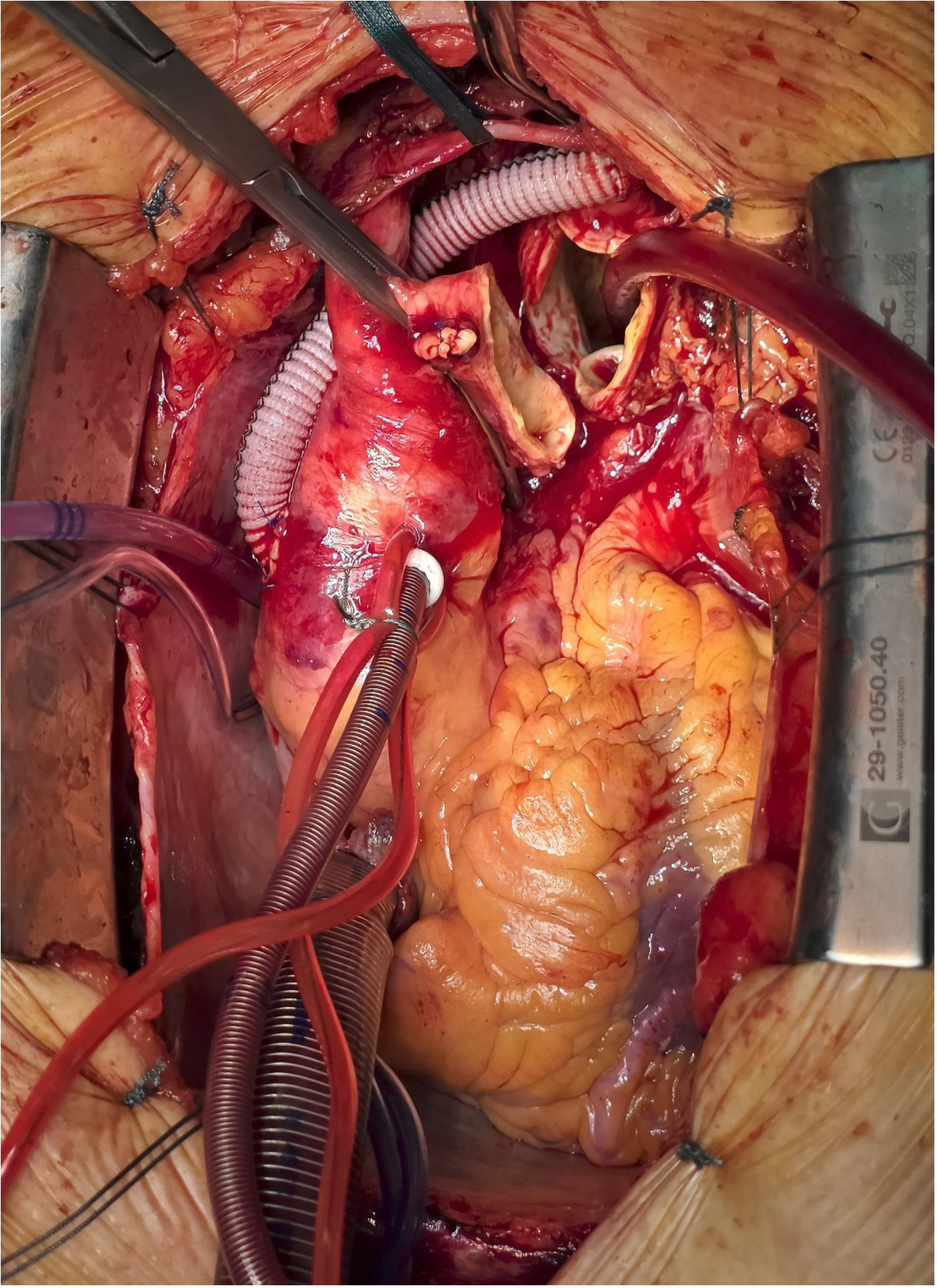
Intraoperative photograph illustrating Variant 2: direct aortic cannulation with extra-anatomical left carotid bypass. The ascending aorta is directly cannulated for cardiopulmonary bypass inflow. An extra-anatomical Dacron side-graft, sutured end-to-side to the proximal ascending aorta and anastomosed to the left common carotid artery, provides uninterrupted simultaneous cerebral and cardiac perfusion during arch repair without a second arterial access site. This variant was employed in patients with aortic pathology extending to distal Zone 2, in whom the proximal ascending aorta remained anatomically suitable for direct cannulation.

Prior to initiation of circulatory arrest, a separate Dacron side-graft was sutured end-to-side to the proximal ascending aorta and anastomosed to the left common carotid artery, establishing an extra-anatomical bypass that ensured uninterrupted cerebral perfusion during the period of arch reconstruction. NIRS monitoring and left ventricular venting were applied in the same manner. SACP via the axillary artery was not used in this variant; bilateral cerebral perfusion was provided via the direct aortic inflow to the right hemispheric vessels and the extra-anatomical left carotid bypass to the left hemispheric.

### Outcome Definitions and Data Collection

The primary outcomes of interest were the feasibility and safety of the beating-heart technique, assessed by freedom from intraoperative conversion to cardioplegic arrest, absence of perioperative myocardial infarction (MI), and absence of low cardiac output syndrome (LCOS). MI was defined as new Q-wave formation or ST-segment changes on postoperative electrocardiography combined with a CK-MB/CK ratio exceeding 10%. LCOS was defined as the requirement for inotropic support (dobutamine >5 μg/kg/min or epinephrine >0.1 μg/kg/min) for more than 24 hours postoperatively, or the need for mechanical circulatory support.

Secondary outcomes included 30-day (in-hospital) mortality, disabling stroke (new permanent neurological deficit at hospital discharge, the Modified Rankin Scale, >2 mRS), postoperative delirium (clinically diagnosed delirium requiring pharmacological treatment), renal replacement therapy, reintubation, tracheotomy, reoperation for bleeding, and extracorporeal life support (ECLS). Biochemical markers of myocardial injury and systemic organ perfusion were recorded prospectively: peak creatine kinase myocardial band (CK-MB) within 48 hours postoperatively, peak intraoperative lactate, and peak lactate within the first 24 hours after surgery. Follow-up data were obtained from outpatient clinic records.

All data were extracted from a prospectively maintained institutional database. Continuous variables are presented as mean ± standard deviation (SD) or median with interquartile range (IQR), as appropriate. Categorical variables are expressed as absolute numbers and percentages. Given the single-arm, proof-of-concept nature of this study, no formal inferential statistical comparisons were performed. All analyses were conducted using Stata (StataNow 19.5, MP, College Station, Texas, USA).

## Results

### Patient Cohort and Baseline Characteristics

Between November 2017 and January 2026, 29 consecutive patients underwent complex aortic arch repair using the total beating-heart technique at our institution. Baseline characteristics are summarised in Table 1. The cohort comprised 19 men (65.5%) and 10 women, with a mean age of 55.4 ± 13.6 years (range 13–71 years). Seven patients (24.1%) had undergone prior surgery or endovascular intervention. Hypertension was the most prevalent cardiovascular risk factor, present in 18 patients (62.1%). Mean preoperative creatinine was 1.0 ± 0.4 mg/dL, reflecting predominantly preserved renal function at baseline.

**Table 1.**
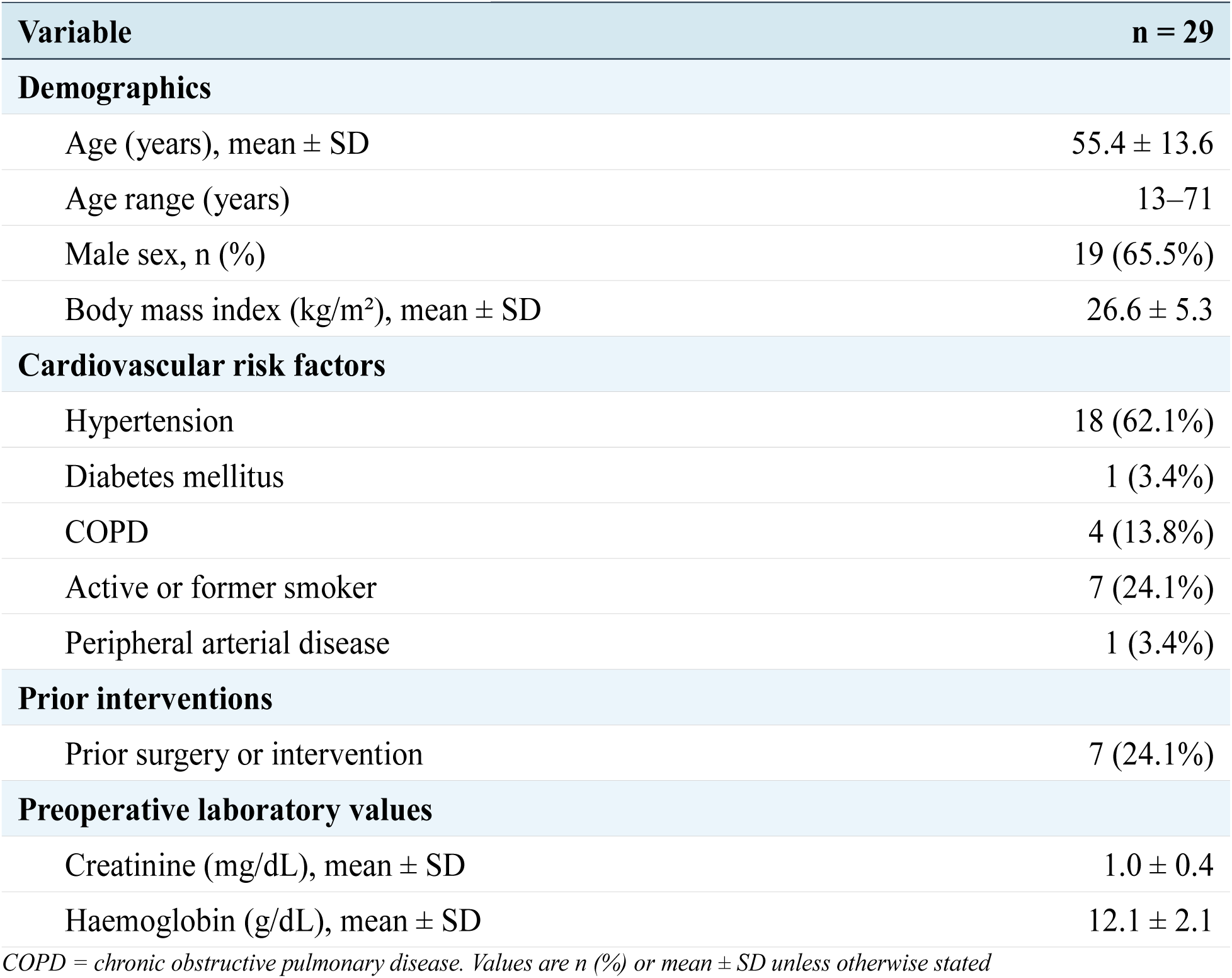
Baseline characteristics.

| Variable | n = 29 |
| --- | --- |
| **Demographics** |  |
| Age (years), mean ± SD | 55.4 ± 13.6 |
| Age range (years) | 13–71 |
| Male sex, n (%) | 19 (65.5%) |
| Body mass index (kg/m<sup>2</sup>), mean ± SD | 26.6 ± 5.3 |
| **Cardiovascular risk factors** |  |
| Hypertension | 18 (62.1%) |
| Diabetes mellitus | 1 (3.4%) |
| COPD | 4 (13.8%) |
| Active or former smoker | 7 (24.1%) |
| Peripheral arterial disease | 1 (3.4%) |
| **Prior interventions** |  |
| Prior surgery or intervention | 7 (24.1%) |
| **Preoperative laboratory values** |  |
| Creatinine (mg/dL), mean ± SD | 1.0 ± 0.4 |
| Haemoglobin (g/dL), mean ± SD | 12.1 ± 2.1 |

*COPD = chronic obstructive pulmonary disease. Values are n (%) or mean ± SD unless otherwise stated*

### Surgical Indications

The spectrum of underlying aortic pathology is detailed in Table 2. Acute aortic dissection accounted for 12 procedures (41.4%), comprising eight cases of acute type B dissection (27.6%) and four cases of acute non-A non-B dissection (13.8%), all performed on an urgent or acute/subacute basis. Arch or thoracic aortic aneurysm was the indication in seven patients (24.1%), and five patients (17.2%) were treated for post-dissection aneurysm or type Ia endoleak following prior thoracic endovascular aortic repair. The remaining four patients (13.8%) presented with miscellaneous pathologies including penetrating aortic ulcer, infectious aortitis with pseudoaneurysm, congenital pseudoaneurysm, and prosthetic graft infection. This breadth of indications reflects the applicability of the technique across a wide range of aortic arch pathologies.

**Table 2.** Surgical indications.

| Indication / Diagnosis | n = 29 |
| --- | --- |
| Acute type B aortic dissection | 8 (27.6%) |
| Acute non-A non-B aortic dissection | 4 (13.8%) |
| Arch or thoracic aortic aneurysm | 7 (24.1%) |
| Post-dissection aneurysm / type Ia TEVAR endoleak | 5 (17.2%) |
| Thoraco-abdominal aortic aneurysm | 1 (3.4%) |
| Other (PAU, aortitis, pseudoaneurysm, prosthetic infection) | 4 (13.8%) |
| **Acute or urgent presentation (dissections)** | **12 (41.4%)** |

PAU = penetrating aortic ulcer; TEVAR = thoracic endovascular aortic repair. Acute/urgent presentation defined as surgery performed for acute aortic dissection (type B or non-A non-B).

### Operative Data and Perfusion Parameters

Operative and perfusion details are presented in Table 3. Total arch replacement was performed in 20 patients (69.0%) and subtotal/partial arch replacement in eight (27.6%). A frozen elephant trunk prosthesis (Thoraflex Hybrid™) was implanted in 16 patients (55.2%). Concomitant coronary artery bypass grafting was performed in two patients (6.9%), both in the early phase of the series.

**Table 3.** Operative and perfusion data.

| Variable | n = 29 |
| --- | --- |
| **Extent of arch repair** |  |
| Total arch replacement | 20 (69.0%) |
| Subtotal/partial arch replacement | 8 (27.6%) |
| Frozen elephant trunk (Thoraflex Hybrid™) | 16 (55.2%) |
| Concomitant CABG | 2 (6.9%) |
| **Perfusion strategy** |  |
| Variant 1 — axillary/subclavian cannulation with needle-vent myocardial perfusion | 24 (82.8%) |
| Variant 2 — direct aortic cannulation with extra-anatomical left carotid bypass | 5 (17.2%) |
| Cardioplegic arrest applied | None (0%) |
| Myocardial perfusion mode | Continuous antegrade warm blood |
| Myocardial perfusion temperature (°C) | 34 |
| Myocardial perfusion flow (mL/min) | 300–400 |
| Target myocardial perfusion pressure (mmHg) | 60–80 |
| Cerebral protection | Selective antegrade (SACP) in all |
| **Perfusion times and temperature** |  |
| Total operative time (min), mean ± SD | 328.3 ± 93.6 |
| Cardiopulmonary bypass time (min), mean ± SD | 193.9 ± 62.9 |
| Circulatory arrest time (min), mean ± SD | 41.3 ± 19.4 |
| Aortic cross-clamp time (min) | 0 — none applied |
| Nadir core temperature (°C), mean ± SD | 29.3 ± 1.5 |

SACP = selective antegrade cerebral perfusion; CABG = coronary artery bypass grafting; FET = frozen elephant trunk. Variant 1: arterial cannulation via the right axillary or subclavian artery, with myocardial perfusion via needle vent in the aortic root (n = 24). Variant 2: direct ascending aortic cannulation with an extra-anatomical side-graft bypass to the left common carotid artery, employed for pathology extending to distal Zone 2 (n = 5). No aortic cross-clamp was applied in any patient. Circulatory arrest denotes the period of reduced lower-body perfusion during arch reconstruction under mild hypothermia.

Variant 1 (axillary cannulation with needle-vent myocardial perfusion) was employed in 24 patients (82.8%), and Variant 2 (direct ascending aortic cannulation with extra-anatomical left carotid bypass) in five patients (17.2%). None of the patients required intraoperative conversion to cardioplegic arrest. Mean CPB time was 193.9 ± 62.9 minutes, mean circulatory arrest time was 41.3 ± 19.4 minutes, and mean total operative time was 328.3 ± 93.6 minutes. Mild hypothermic circulatory arrest was employed uniformly, with a mean nadir core temperature of 29.3 ± 1.5°C. Continuous antegrade myocardial perfusion was maintained throughout each procedure at 34°C and 300–400 mL/min, with myocardial perfusion pressure continuously monitored and maintained at a target of 60–80 mmHg.

### Cardiac Outcomes

Cardiac outcomes are reported in Table 4. No patient sustained a perioperative myocardial infarction. Low cardiac output syndrome did not occur in any patient. These findings represent the primary outcome of this proof-of-concept study and support the hypothesis that uninterrupted coronary perfusion during aortic arch repair may effectively prevent clinically significant myocardial ischaemic injury, even in the context of complex, prolonged, and high-risk reconstructions.

**Table 4.**
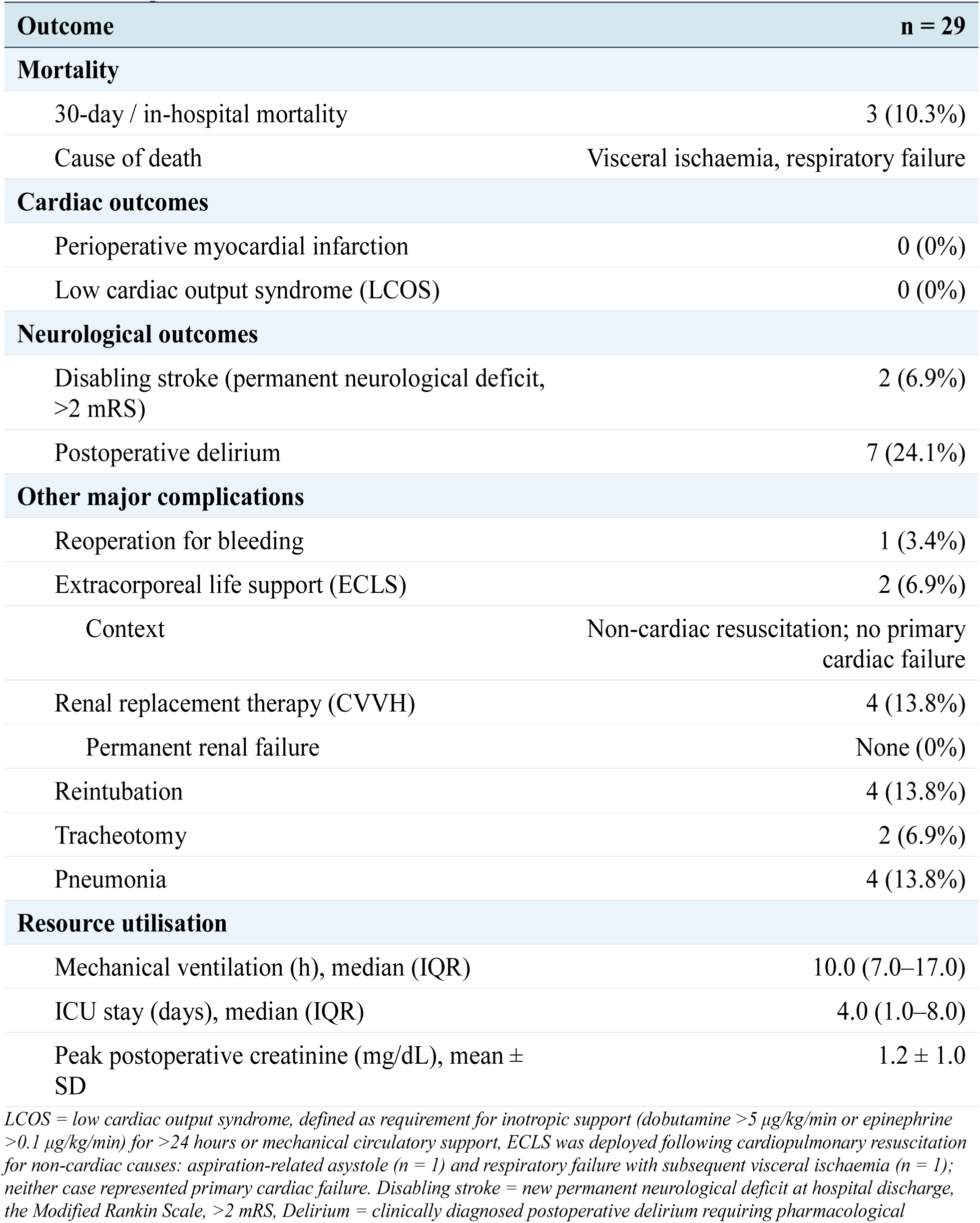
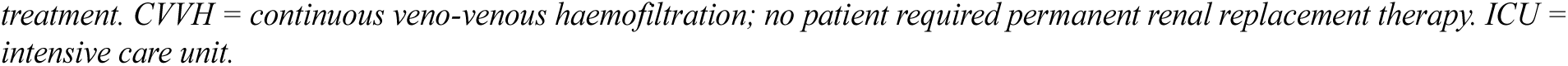
In-hospital outcomes.

| Outcome | n = 29 |
| --- | --- |
| Mortality |  |
| 30-day / in-hospital mortality | 3 (10.3%) |
| Cause of death | Visceral ischaemia, respiratory failure |
| Cardiac outcomes |  |
| Perioperative myocardial infarction | 0 (0%) |
| Low cardiac output syndrome (LCOS) | 0 (0%) |
| Neurological outcomes |  |
| Disabling stroke (permanent neurological deficit, >2 mRS) | 2 (6.9%) |
| Postoperative delirium | 7 (24.1%) |
| Other major complications |  |
| Reoperation for bleeding | 1 (3.4%) |
| Extracorporeal life support (ECLS) | 2 (6.9%) |
| Context | Non-cardiac resuscitation; no primary cardiac failure |
| Renal replacement therapy (CVVH) | 4 (13.8%) |
| Permanent renal failure | None (0%) |
| Reintubation | 4 (13.8%) |
| Tracheotomy | 2 (6.9%) |
| Pneumonia | 4 (13.8%) |
| Resource utilisation |  |
| Mechanical ventilation (h), median (IQR) | 10.0 (7.0–17.0) |
| ICU stay (days), median (IQR) | 4.0 (1.0–8.0) |
| Peak postoperative creatinine (mg/dL), mean ± SD | 1.2 ± 1.0 |

LCOS = low cardiac output syndrome, defined as requirement for inotropic support (dobutamine >5 μg/kg/min or epinephrine >0.1 μg/kg/min) for >24 hours or mechanical circulatory support, ECLS was deployed following cardiopulmonary resuscitation for non-cardiac causes: aspiration-related asystole (n = 1) and respiratory failure with subsequent visceral ischaemia (n = 1); neither case represented primary cardiac failure. Disabling stroke = new permanent neurological deficit at hospital discharge, the Modified Rankin Scale, >2 mRS, Delirium = clinically diagnosed postoperative delirium requiring pharmacological
treatment. CVVH = continuous veno-venous haemofiltration; no patient required permanent renal replacement therapy. ICU = intensive care unit.

Two patients (6.9%) required in postoperative course extracorporeal life support (ECLS); however, in neither case did this represent primary myocardial failure. The first patient developed aspiration-related respiratory compromise with subsequent cardiac asystole; the second experienced progressive respiratory failure with visceral ischaemia identified as the underlying aetiology. Both events arose from non-cardiac primary organ dysfunction, and ECLS was deployed as a rescue measure following cardiopulmonary resuscitation.

### Biomarkers of Myocardial and Systemic Organ Injury

Biomarker data are summarised in Table 5. The median peak CK-MB was 44.0 U/L (IQR 34.0–59.0; mean 50.1 ± 26.6 U/L). No patient exceeded the electrocardiographic or biochemical threshold criteria for perioperative myocardial infarction. These values are consistent with the myocardial enzyme release attributable to surgical manipulation and the physiological demands of CPB, rather than ischaemic necrosis, and corroborate the clinical absence of low cardiac output syndrome across the cohort. Mean peak intraoperative lactate was 4.9 ± 2.1 mmol/L, reflecting the metabolic demands of hypothermic circulatory arrest and the period of reduced lower-body perfusion during arch reconstruction. Mean peak lactate declined to 2.9 ± 1.2 mmol/L within the first 24 hours postoperatively, indicating prompt metabolic recovery and adequate global organ perfusion following re-establishment of full circulatory flow.

**Table 5.** Biomarkers of myocardial and systemic organ injury.

| **Biomarker** | **n = 29** |
| --- | --- |
| **Myocardial injury marker** |  |
| Peak CK-MB (U/L), mean ± SD | 50.1 ± 26.6 |
| Peak CK-MB (U/L), median (IQR) | 44.0 (34.0–59.0) |
| **Global organ perfusion markers** |  |
| Peak intraoperative lactate (mmol/L), mean ± SD | 4.9 ± 2.1 |
| Peak 24h postoperative lactate (mmol/L), mean ± SD | 2.9 ± 1.2 |

CK-MB = creatine kinase myocardial band. Peak CK-MB: highest value within 48h postoperatively. Lactate values represent highest recorded intraoperative value and highest value within the first 24h postoperatively, respectively. No patient met electrocardiographic or biochemical criteria for perioperative myocardial infarction.

### Neurological Outcomes

Two patients (6.9%) sustained a disabling stroke, defined as a new permanent neurological deficit at hospital discharge, with a grade of at least moderate disability in The Modified Rankin Scale (>2 mRS). Postoperative delirium requiring pharmacological treatment occurred in seven patients (24.1%). This rate is consistent with contemporary series of complex aortic arch surgery and reflects the high proportion of acute dissections and haemodynamically compromised presentations within this cohort, rather than a complication specific to the perfusion strategy.

### Postoperative Morbidity and Resource Utilisation

Reoperation for bleeding was required in one patient (3.4%). Renal replacement therapy in the form of continuous veno-venous haemofiltration (CVVH) was necessary in four patients (13.8%); no patient required permanent renal replacement therapy at discharge. Respiratory complications included reintubation in four patients (13.8%), tracheotomy in two (6.9%), and pneumonia in four (13.8%). Median mechanical ventilation time was 10.0 hours (IQR 7.0–17.0 hours) and median intensive care unit stay was 4.0 days (IQR 1.0–8.0 days).

### Thirty-Day Mortality

Thirty-day mortality was 10.3% (n = 3). These three deaths were attributable to aspiration-related respiratory compromise or to visceral ischaemia arising in the context of acute type B aortic dissection — a pathological entity characterised by dynamic malperfusion and a historically elevated operative mortality irrespective of the myocardial protection strategy employed. No death was attributable to primary cardiac failure, perioperative myocardial infarction, or any complication directly related to the beating-heart perfusion technique. There were no intraoperative deaths.

### Follow-Up

Follow-up data are presented in Table 6. Median follow-up was 6.0 months (IQR 1.0–33.0 months; mean 19.3 ± 25.5 months; range 0–83 months). Among the 26 patients who survived to hospital discharge, three (11.5%) died during follow-up. All late deaths were attributable to progressive distal aortic pathology — including aortic expansion and dissection-related complications in the downstream thoraco-abdominal aorta — and none was directly attributable to the index arch procedure or the myocardial protection strategy. Survival among 30-day survivors at the latest available follow-up was 88.5% (23 of 26 patients).

**Table 6.** Follow-up.

| **Variable** | **n = 29** |
| --- | --- |
| **Follow-up duration** |  |
| Mean ± SD (months) | 19.3 ± 25.5 |
| Median (IQR) (months) | 6.0 (1.0–33.0) |
| Range (months) | 0–83 |
| **Late outcomes** |  |
| Late deaths, n (%) | 3 (11.5% of 30-day survivors) |
| Cause of late death | Progressive distal aortic pathology (all 3) |
| Survival at latest follow-up (30-day survivors) | 23/26 (88.5%) |

Follow-up obtained via outpatient clinic visits and cross-sectional imaging review. All late deaths were due to progressive distal aortic pathology and were not attributable to the index procedure or the myocardial protection strategy.

## Discussion

### The Problem of Myocardial Ischaemia in Complex Arch Surgery

Complex aortic arch repair is among the most physiologically demanding operations in cardiac surgery. Beyond the challenges of cerebral and visceral protection during circulatory arrest, the heart itself is subjected to a protracted ischaemic injury that is often underappreciated in the literature. In most contemporary series of total arch replacement, the cardiac ischaemia time — defined as the duration of cardioplegic arrest — routinely exceeds 90 to 120 minutes; in cases requiring concomitant aortic root reconstruction, coronary reimplantation, or valve surgery, cumulative arrest times approaching or exceeding 150 minutes are not uncommon(1,2,9–11). Even with meticulous intermittent cold blood cardioplegia, such durations are associated with progressive myocardial energy depletion, intracellular calcium accumulation, and impaired contractile recovery — a syndrome that may, in its clinical form, manifest as low cardiac output, prolonged vasopressor and inotrope dependency, and, at its most severe, multiorgan failure.

That cardiac failure is consistently underreported in overviews of aortic arch surgery reflects a well-recognised ascertainment and reporting bias rather than a genuine absence of the complication. In the landmark series by Martens et al., cardiac failure accounted for 9% of 30-day deaths in the cardioplegia-alone group, and multivariate analysis identified CMP as an independent predictor of reduced early mortality and LCOS — directly implicating prolonged myocardial ischaemia as a modifiable driver of perioperative death in complex arch surgery(7). Shrestha et al., reporting thirty years of institutional experience, identified cardiac causes as a major contributor to perioperative mortality in complex arch cases(2). This pattern is consistent with the well-established association between prolonged cardiac ischaemia time and postoperative ventricular dysfunction in all forms of cardiac surgery(12).

These observations provide the physiological rationale for exploring myocardial protection strategies that eliminate or minimise the period of global coronary ischaemia during arch repair. Martens and colleagues were the first to systematically demonstrate in a clinical series, that non-cardioplegic continuous myocardial perfusion (CMP) — initiated after an initial period of cardioplegic arrest to complete any necessary cardiac procedures — significantly reduced cardiac ischaemia time (49 ± 32 versus 149 ± 56 minutes), LCOS (3% versus 22%), and 30-day mortality (6% versus 21%) compared with cardioplegia alone(7). Their landmark study established that the heart does not need to remain arrested throughout the entirety of arch repair. The concept of continuous myocardial perfusion has since then been adopted in multiple centres — including ours. What remained unanswered was whether cardioplegic arrest is necessary at any stage of the procedure — that is, whether the entire operation can be performed on a continuously and uninterruptedly perfused heart.

### A Conceptually Distinct Strategy: Eliminating Cardioplegic Arrest Entirely

The technique described in the present series differs from all previously reported CMP strategies in one defining respect: cardioplegic arrest was not applied at any stage of the procedure in any patient. In the series of Martens et al., a period of cardioplegic arrest was employed to complete intracardiac and aortic root procedures before CMP was initiated; the transition from arrest to continuous perfusion was a deliberate midprocedural step, not an alternative to arrest(7). Similarly, the paediatric beating-heart protocols reported by Ruffer and colleagues and by Kotani et al. relied on initial cardioplegic induction in the majority of cases(8,13). In the isolated adult case reports of Abu-Omar et al. and Di Natale et al., temporary cold cardiac arrest preceded the institution of myocardial perfusion(14,15). In every previously published series, a period of deliberate global myocardial ischaemia was therefore imposed at some point during the procedure.

In the present cohort, by contrast, the ascending aorta was cross-clamped and continuous antegrade myocardial perfusion via the aortic root needle vent was established immediately and maintained without interruption throughout. The heart was never chemically or electrically arrested; coronary perfusion was never deliberately interrupted; global myocardial ischaemia was never induced. Our technique is based on a warm, pressure-monitored, flow-controlled myocardial perfusion system that operates throughout the entire procedure, from initial clamping of the ascending aorta to weaning from cardiopulmonary bypass, without any preceding period of arrest.

This conceptual distinction has direct physiological implications. By eliminating the initial cardioplegic insult entirely, the approach avoids the ischaemia-reperfusion injury that accompanies even optimally managed cardiac arrest, preserves mitochondrial energetics and calcium homeostasis from the outset, and maintains continuous — if hypothermically attenuated — myocardial perfusion throughout. The anatomical prerequisites for this strategy are deliberately restrictive: a macroscopically normal aortic root and ascending aorta are required to seat the perfusion needle vent securely and deliver unobstructed coronary perfusion, and significant coronary artery disease constitutes an absolute contraindication, as regional ischaemia may persist despite adequate aortic root perfusion pressure in the presence of haemodynamically significant coronary stenoses. These constraints define a specific but clinically relevant patient subset — those with pathology confined to the arch and descending aorta while the proximal aortic and coronary anatomy remain preserved — encompassing the majority of acute type B and non-A non-B dissections, isolated arch aneurysms, and reoperative arch pathology following prior endovascular treatment.

### Contextualisation of Outcomes Within the Contemporary Literature

The primary cardiac outcome of this series — no perioperative myocardial infarction and no low cardiac output syndrome across 29 patients operated on under continuous myocardial perfusion — constitutes the central finding of this proof-of-concept study. These results should be interpreted in the context of the prevailing surgical benchmarks. In the FET series of Shrestha et al. reporting 100 consecutive FET implantations, mean cardiac ischaemia time was 101 ± 65 minutes, and perioperative mortality was 7%.(2) In a contemporary single-centre experience of 132 patients undergoing total arch replacement with FET for mixed aortic pathology, median cardiac ischaemia time was 89 minutes and 30-day mortality was 7.6%(5). Notably, in the cardioplegia-alone arm of the Martens series — the largest systematic comparison of myocardial protection strategies in arch surgery to date — LCOS occurred in 22% of patients and cardiac causes accounted for 9% of operative deaths, directly implicating prolonged myocardial ischaemia as a modifiable driver of perioperative mortality in this setting(7). Against this backdrop, the complete absence of LCOS in our cohort — despite a mean CPB time of 194 minutes and a procedure complexity reflected in the high proportion of total arch replacements (69%) and FET implantations (55%) — provides a coherent and biologically plausible signal in favour of the no-arrest strategy.

Biomarker data corroborate this interpretation. In the institutional control cohort of 64 patients undergoing elective total or subtotal arch replacement with conventional cardioplegia at the same centre over a contemporaneous period, mean peak CK-MB was 73.0 ± 59.1 U/L — significantly higher than the 50.1 ± 26.6 U/L observed in the beating-heart group. The lower absolute values and markedly reduced CK-MB in the beating-heart cohort (SD 26.6 versus 59.1 U/L) suggest both a lower level of myocardial injury and a more consistent degree of myocardial protection across patients of varying complexity. These figures are broadly consistent with those reported by Martens et al., in whose series the cardioplegia-alone group had mean peak CK-MB of 102 ± 106 U/L versus 82 ± 65 U/L in the CMP group — itself a strategy that still incorporated initial cardioplegic arrest(7). The further reduction in CK-MB observed in our series is consistent with the hypothesis that the degree of myocardial injury correlates incrementally with the duration and completeness of global coronary ischaemia.

Thirty-day mortality of 10.3% (3 of 29 patients) warrants careful contextualisation. These three deaths occurred mostly in patients operated on for acute type B aortic dissection complicated by visceral malperfusion — a pathological entity for which open surgical repair carries a pooled 30-day mortality of approximately 19% in systematic analyses of the published literature(16). No death in this series was attributable to primary cardiac failure, which stands in contrast to the historical experience in comparable cohorts where cardiac causes consistently contribute to operative mortality(7). The disabling strokes (6.9%) and the postoperative delirium rate (24.1%) are consistent with published benchmarks for complex arch surgery in a cohort enriched for acute dissection, urgent presentation, and prior aortic intervention(5,11).

In the institutional control cohort, LCO was observed in 8 of 64 patients (12.5%) and 30-day mortality was 7.8%, consistent with the published literature for elective arch repair(5). The lower 30-day mortality in the control group compared with the beating-heart cohort (7.8% versus 10.3%) reflects the substantially different risk profiles of the two groups: the control cohort consisted exclusively of elective cases without aortic dissection, while the beating-heart series included 41% acute dissections with malperfusion — the highest-risk indication for arch surgery. A formal comparative analysis of outcomes between the two groups was not performed, as the baseline differences in indication, urgency, and patient selection preclude any meaningful inference of treatment effect from an unadjusted comparison.

### Comparison With the Contemporary Institutional Control Cohort

To contextualise the outcomes of the beating-heart series within the institutional experience, we analysed a contemporaneous cohort of 64 patients who underwent elective total or subtotal aortic arch replacement with conventional cardioplegia at the same centre over the same calendar period (Table 7). These two groups differed substantially in baseline characteristics and operative profile, precluding causal inference; the analysis is presented as descriptive and hypothesis-generating rather than confirmatory.

**Table 7.**
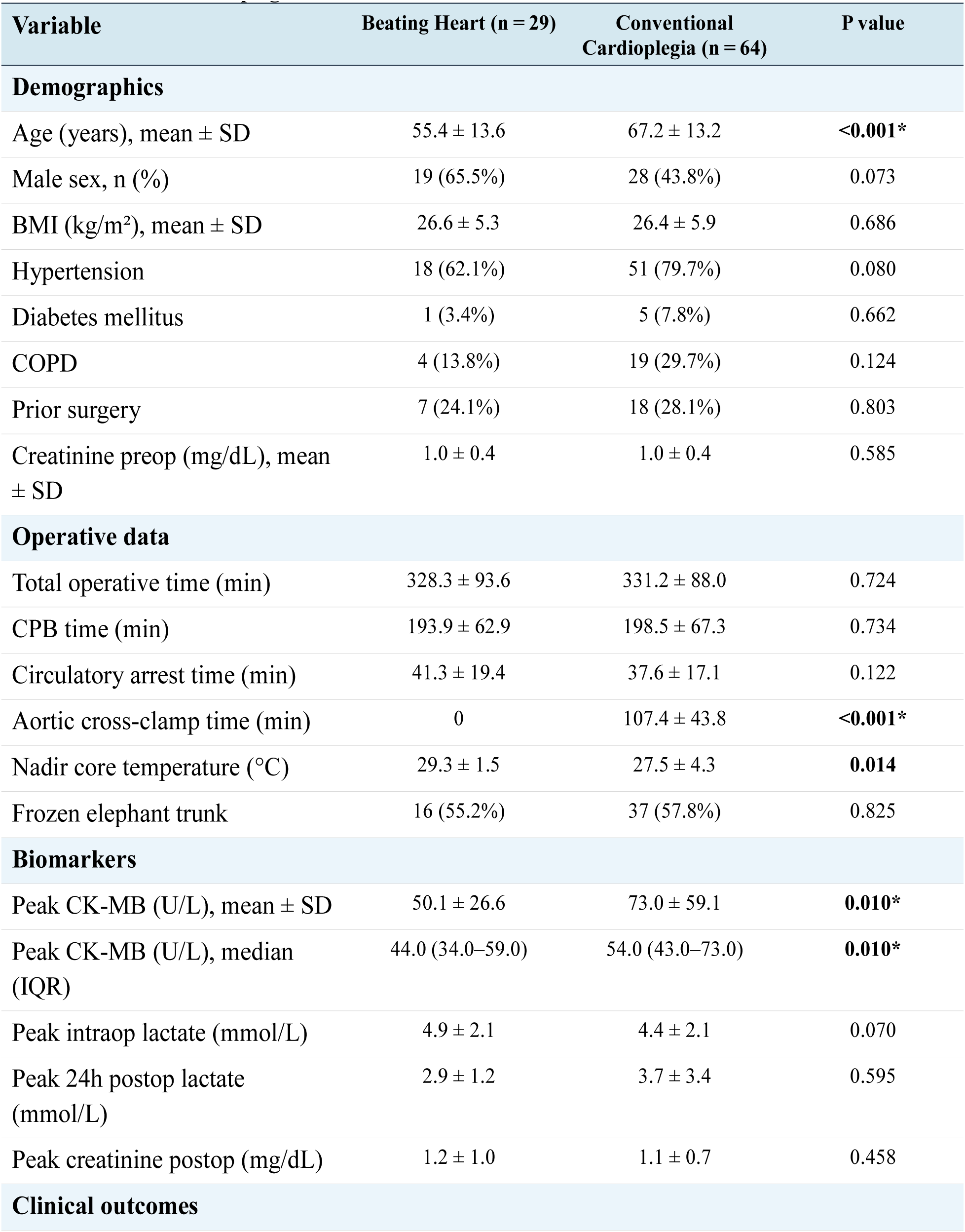
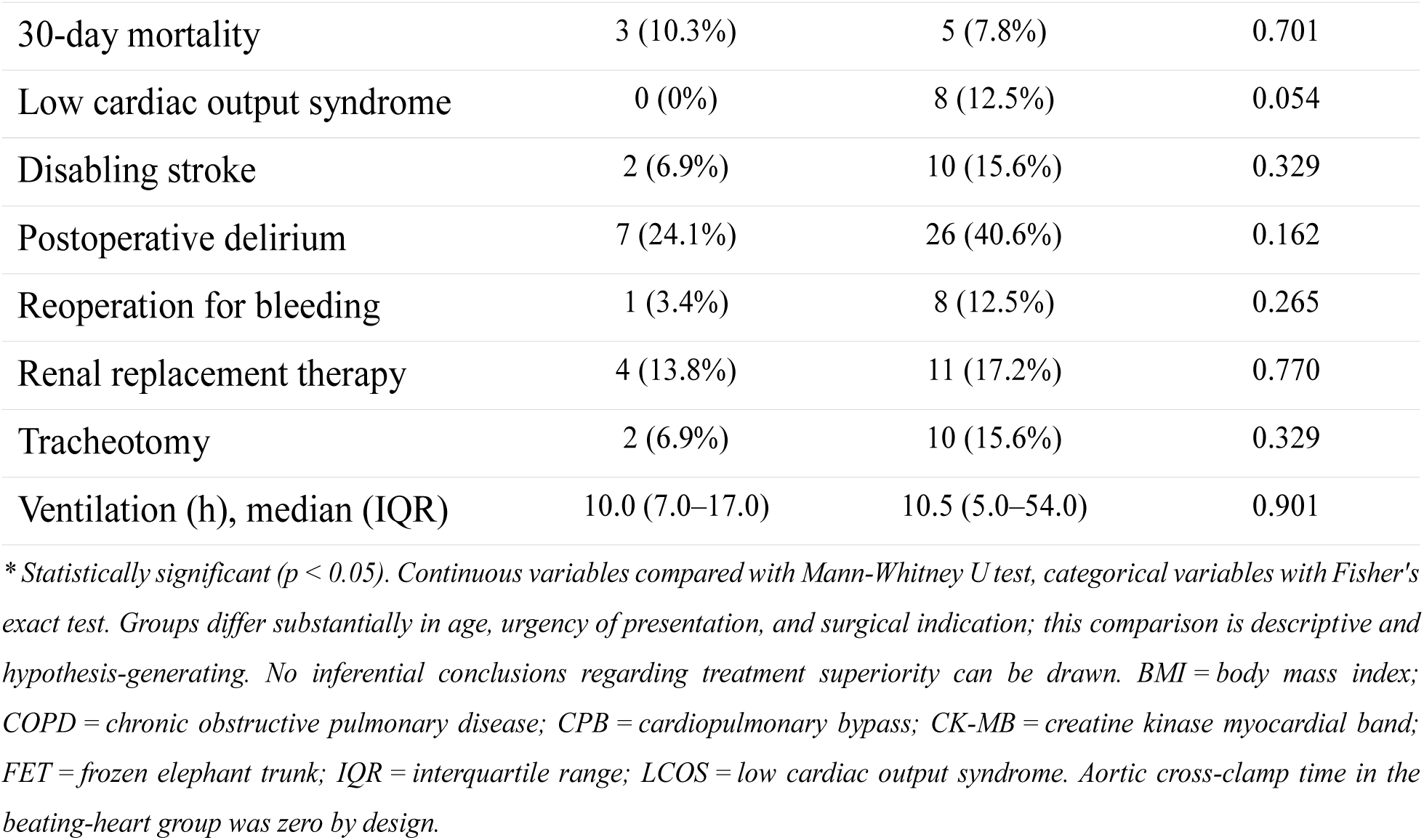
Comparative analysis: beating-heart technique versus contemporary institutional cohort with conventional cardioplegia.

| Variable | Beating Heart (n = 29) | Conventional Cardioplegia (n = 64) | P value |
| --- | --- | --- | --- |
| **Demographics** |  |  |  |
| Age (years), mean ± SD | 55.4 ± 13.6 | 67.2 ± 13.2 | **<0.001\*** |
| Male sex, n (%) | 19 (65.5%) | 28 (43.8%) | 0.073 |
| BMI (kg/m<sup>2</sup>), mean ± SD | 26.6 ± 5.3 | 26.4 ± 5.9 | 0.686 |
| Hypertension | 18 (62.1%) | 51 (79.7%) | 0.080 |
| Diabetes mellitus | 1 (3.4%) | 5 (7.8%) | 0.662 |
| COPD | 4 (13.8%) | 19 (29.7%) | 0.124 |
| Prior surgery | 7 (24.1%) | 18 (28.1%) | 0.803 |
| Creatinine preop (mg/dL), mean ± SD | 1.0 ± 0.4 | 1.0 ± 0.4 | 0.585 |
| **Operative data** |  |  |  |
| Total operative time (min) | 328.3 ± 93.6 | 331.2 ± 88.0 | 0.724 |
| CPB time (min) | 193.9 ± 62.9 | 198.5 ± 67.3 | 0.734 |
| Circulatory arrest time (min) | 41.3 ± 19.4 | 37.6 ± 17.1 | 0.122 |
| Aortic cross-clamp time (min) | 0 | 107.4 ± 43.8 | **<0.001\*** |
| Nadir core temperature (°C) | 29.3 ± 1.5 | 27.5 ± 4.3 | **0.014** |
| Frozen elephant trunk | 16 (55.2%) | 37 (57.8%) | 0.825 |
| **Biomarkers** |  |  |  |
| Peak CK-MB (U/L), mean ± SD | 50.1 ± 26.6 | 73.0 ± 59.1 | **0.010\*** |
| Peak CK-MB (U/L), median (IQR) | 44.0 (34.0–59.0) | 54.0 (43.0–73.0) | **0.010\*** |
| Peak intraop lactate (mmol/L) | 4.9 ± 2.1 | 4.4 ± 2.1 | 0.070 |
| Peak 24h postop lactate (mmol/L) | 2.9 ± 1.2 | 3.7 ± 3.4 | 0.595 |
| Peak creatinine postop (mg/dL) | 1.2 ± 1.0 | 1.1 ± 0.7 | 0.458 |
| **Clinical outcomes** |  |  |  |
| 30-day mortality | 3 (10.3%) | 5 (7.8%) | 0.701 |
| Low cardiac output syndrome | 0 (0%) | 8 (12.5%) | 0.054 |
| Disabling stroke | 2 (6.9%) | 10 (15.6%) | 0.329 |
| Postoperative delirium | 7 (24.1%) | 26 (40.6%) | 0.162 |
| Reoperation for bleeding | 1 (3.4%) | 8 (12.5%) | 0.265 |
| Renal replacement therapy | 4 (13.8%) | 11 (17.2%) | 0.770 |
| Tracheotomy | 2 (6.9%) | 10 (15.6%) | 0.329 |
| Ventilation (h), median (IQR) | 10.0 (7.0–17.0) | 10.5 (5.0–54.0) | 0.901 |

\* Statistically significant ( $p < 0.05$ ). Continuous variables compared with Mann-Whitney U test, categorical variables with Fisher's exact test. Groups differ substantially in age, urgency of presentation, and surgical indication; this comparison is descriptive and hypothesis-generating. No inferential conclusions regarding treatment superiority can be drawn. BMI = body mass index; COPD = chronic obstructive pulmonary disease; CPB = cardiopulmonary bypass; CK-MB = creatine kinase myocardial band; FET = frozen elephant trunk; IQR = interquartile range; LCOS = low cardiac output syndrome. Aortic cross-clamp time in the beating-heart group was zero by design.

The most pronounced demographic difference was age: patients in the beating-heart group were a mean of 11.8 years younger (55.4 ± 13.6 versus 67.2 ± 13.2 years; p<0.001), reflecting the different pathological substrate — predominantly dissections in the beating-heart series versus elective degenerative aneurysm in the control cohort. Critically, however, operative complexity as measured by perfusion duration was indistinguishable between groups: total operative time (328 ± 94 versus 331 ± 88 min; p = 0.724), CPB time (194 ± 63 versus 199 ± 67 min; p = 0.734), and circulatory arrest time (41 ± 19 versus 38 ± 17 min; p = 0.122) were comparable. The proportion of patients receiving a frozen elephant trunk prosthesis was similarly matched between groups (55.2% versus 57.8%; p = 0.825), confirming that the two cohorts underwent procedures of equivalent technical complexity.

The sole statistically significant difference in biomarkers was peak CK-MB, which was significantly lower in the beating-heart group (mean 50.1 ± 26.6 versus 73.0 ± 59.1 U/L; p = 0.010). The standard deviation of CK-MB was more than twice as wide in the control group (59.1 versus 26.6 U/L), reflecting greater inter-individual variability in myocardial injury under conditions of variable-duration ischaemia — a heterogeneity that is markedly attenuated when protection is provided by uninterrupted coronary perfusion. Peak 24-hour postoperative lactate showed a numerical trend favouring the beating-heart group (2.9 ± 1.2 versus 3.7 ± 3.4 mmol/L; p = 0.595), without reaching statistical significance, likely due to the greater metabolic heterogeneity of the control cohort.

Regarding clinical outcomes, low cardiac output syndrome occurred in 8 of 64 control patients (12.5%) compared with zero in the beating-heart group (p = 0.054). While this difference did not reach conventional statistical significance — attributable to the modest sample size of the beating-heart series — the absolute risk difference of 12.5 percentage points is clinically substantial, and the consistent direction of effect across all cardiac outcome measures (LCOS, CK-MB, perioperative MI) constitutes a coherent and biologically plausible signal in support of the myocardial protection hypothesis. Thirty-day mortality was 10.3% versus 7.8% (p = 0.701), a non-significant difference explained by the higher-risk cohort of the beating-heart cohort: all three deaths occurred in patients with acute type B dissection complicated by visceral malperfusion, whereas the control cohort consisted exclusively of elective cases. No death in the beating-heart group was attributable to primary cardiac failure. Rates of disabling stroke (6.9% versus 15.6%; p = 0.329), postoperative delirium (24.1% versus 40.6%; p = 0.162), and reoperation for bleeding (3.4% versus 12.5%; p = 0.265) were numerically lower in the beating-heart group across all domains, though none reached individual statistical significance, consistent with the constraints of a proof-of-concept series.

Taken together, these data suggest that at comparable operative complexity — matched by CPB duration, circulatory arrest time, operative time, and FET utilisation — the no-arrest myocardial protection strategy is associated with lower myocardial enzyme release and a consistent trend towards reduced cardiac and non-cardiac morbidity, without any detectable penalty in neurological, renal, or pulmonary outcomes. The confounding effects of age, indication, and urgency cannot be eliminated in an unadjusted comparison of groups as clinically distinct as these, and no causal conclusions regarding superiority can be drawn. What these findings do provide is a sufficiently compelling and internally consistent hypothesis — grounded in biological plausibility and supported by a coherent biomarker and clinical outcome signal — to justify a prospective, adequately powered, multicentre evaluation of the no-arrest approach.

### Limitations

Several important limitations must be acknowledged. This is a single-centre, single-surgeon series of 29 patients accumulated over nearly nine years, and the modest sample size, while sufficient to establish proof of concept and generate an initial safety signal, is inadequate to support definitive conclusions regarding the superiority of the total beating-heart approach over conventional cardioplegia-based strategies, or to permit statistically reliable estimation of differences in low-frequency outcomes such as stroke or renal failure. The technique evolved substantially during the observation period, with progressive refinement of patient selection criteria, perfusion parameters, and the introduction of the two-variant operative strategy; earlier cases — particularly those involving acute dissection and the two patients who required unplanned coronary artery bypass grafting — reflect a phase of technical development rather than a mature, standardised protocol, and outcomes in these cases cannot be attributed entirely to the perfusion strategy. A learning-curve effect is inherent in any single-centre, single-surgeon experience and should be acknowledged when extrapolating these results to other centres or operators.

The absence of a contemporaneous, propensity-matched control group precludes causal inference. The institutional control cohort presented for contextual reference is substantially older, predominantly female, and composed exclusively of elective cases without dissection — a risk profile that differs fundamentally from the beating-heart cohort in both baseline characteristics and operative risk. Formal inferential comparisons under these conditions would generate systematically biased estimates of treatment effect and have accordingly not been performed. This study is presented, and should be interpreted, as a proof-of-concept series establishing feasibility, reporting an initial safety signal, and generating hypotheses for future controlled evaluation — not as evidence of superiority over standard techniques.

Follow-up duration is heterogeneous and, for a substantial proportion of patients, short. The median follow-up of six months precludes meaningful assessment of long-term survival, freedom from reoperation, or the durability of the arch reconstruction. Prospective evaluation with standardised follow-up intervals and imaging protocols will be required to address these questions. Finally, the anatomical prerequisites of the technique — a normal aortic root and the absence of coronary artery disease — restrict its applicability to a defined subset of arch surgery patients, and its extension to those requiring concomitant root replacement or valve surgery in its current form remains technically unsolved.

### Conclusions

Total aortic arch repair performed on the continuously perfused beating heart, without any period of cardioplegic arrest, is technically feasible and clinically safe in appropriately selected patients. In this single-centre proof-of-concept series of 29 patients encompassing a broad spectrum of aortic arch pathologies — including 41% acute or urgent dissections — the complete elimination of global myocardial ischaemia was associated with no perioperative myocardial infarctions, no low cardiac output syndrome, and lower peak CK-MB values than those observed in a contemporaneous institutional cohort undergoing conventional arch repair with cardioplegia. These findings establish the proof-of-concept for a strategy that may further reduce the cardiac morbidity and mortality of complex aortic arch surgery. Prospective, multicentre evaluation with rigorous patient selection, standardised perfusion protocols, and systematic follow-up is required to define the role of this approach within the broader armamentarium of arch organ protection.

## Data Availability

The data underlying this article are not publicly available owing to patient privacy restrictions and institutional data governance policies. De-identified data supporting the findings of this study may be made available upon reasonable request to the corresponding author, subject to approval by the institutional data protection officer and the ethics committee of the University Hospital Münster.

## Acknowledgments

We thank Agata Trojanowska, MSc (https://trojanowska.myportfolio.com/), for providing surgical illustrations.

## Supplemental Material

**Video S1. *Beating-heart aortic arch repair prior to arch reconstruction (Variant 1).*** The video demonstrates continuous cardiac activity during the period of aortic cross-clamping and selective antegrade cerebral perfusion, prior to the aortic arch repair. The ascending aorta is cross-clamped at the sinotubular junction (proximal clamp), continuous antegrade myocardial perfusion is established via the aortic root needle vent, delivering warm oxygenated blood. The heart is seen beating in sinus rhythm, demonstrating preserved myocardial contractile activity in the absence of any cardioplegic arrest. Left ventricular decompression is maintained via a vent in the right superior pulmonary vein. Suction in the open aortic arch is also visible.

**Video S2. *Preserved cardiac function after frozen elephant trunk implantation (Variant 1).*** The video demonstrates sustained myocardial contractile activity following completion of total aortic arch replacement with frozen elephant trunk implantation. Continuous antegrade myocardial perfusion via the aortic root needle vent has been maintained uninterrupted throughout the arch reconstruction. The preserved cardiac function visible in this sequence reflects the absence of global myocardial ischaemia at any point during the procedure.

## List of Abbreviations

CABG: Coronary artery bypass grafting
CK-MB: Creatine kinase myocardial band
CMP: Continuous myocardial perfusion
COPD: Chronic obstructive pulmonary disease
CPB: Cardiopulmonary bypass
CVVH: Continuous veno-venous haemofiltration
ECLS: Extracorporeal life support
FET: Frozen elephant trunk
HCA: Hypothermic circulatory arrest
ICU: Intensive care unit
IQR: Interquartile range
LCOS: Low cardiac output syndrome
LV: Left ventricle
MI: Myocardial infarction
MPF: Myocardial perfusion flow
MPP: Myocardial perfusion pressure
NIRS: Near-infrared spectroscopy
PAU: Penetrating aortic ulcer
RCA: Right coronary artery
RV: Right ventricle
SACP: Selective antegrade cerebral perfusion
SD: Standard deviation
TAAA: Thoraco-abdominal aortic aneurysm
TEVAR: Thoracic endovascular aortic repair

